# An Empirical Study on KDIGO-Defined Acute Kidney Injury Prediction in the Intensive Care Unit

**DOI:** 10.1101/2024.02.01.24302063

**Authors:** Xinrui Lyu, Bowen Fan, Matthias Hüser, Philip Hartout, Thomas Gumbsch, Martin Faltys, Tobias M. Merz, Gunnar Rätsch, Karsten Borgwardt

**Affiliations:** Department of Computer Science, ETH Zürich, Switzerland; NEXUS Personalized Health Technologies, ETH Zürich, Switzerland; Swiss Institute for Bioinformatics, Lausanne, Switzerland; Department of Biosystems Science and Engineering, ETH Zürich, Switzerland; Department of Machine Learning and Systems Biology, Max Planck Institute of Biochemistry, Martinsried, Germany; Department of Intensive Care, Austin Hospital, Melbourne, Australia; Department of Intensive Care Medicine, University Hospital, University of Bern, Switzerland; Cardiovascular Intensive Care Unit, Auckland City Hospital, New Zealand; Medical Informatics Unit, Zürich University Hospital, Switzerland; AI Center at ETH Zürich, Switzerland; Department of Biology, ETH Zürich, Switzerland

## Abstract

**Motivation:** Acute kidney injury (AKI) is a syndrome that affects a large fraction of all critically ill patients, and early diagnosis to receive adequate treatment is as imperative as it is challenging to make early. Consequently, machine learning approaches have been developed to predict AKI ahead of time. However, the prevalence of AKI is often underestimated in state-of-the-art approaches, as they rely on an AKI event annotation solely based on creatinine, ignoring urine output.

**Methods:** We construct and evaluate early warning systems for AKI in a multi-disciplinary ICU setting, using the complete KDIGO definition of AKI. We propose several variants of gradient-boosted decision trees (GBDT)-based models, including a novel time-stacking based approach. A state-of-the-art LSTM-based model previously proposed for AKI prediction is used as a comparison, which was not specifically evaluated in ICU settings yet.

**Results:** We find that optimal performance is achieved by using GBDT with the time-based stacking technique (AUPRC=65.7%, compared with the LSTM-based model’s AUPRC=62.6%), which is motivated by the high relevance of time since ICU admission for this task. Both models show mildly reduced performance in the limited training data setting, perform fairly across different subcohorts, and exhibit no issues in gender transfer.

**Conclusion:** Following the official KDIGO definition substantially increases the number of annotated AKI events. In our study GBDTs outperform LSTM models for AKI prediction. Generally, we find that both model types are robust in a variety of challenging settings arising for ICU data.

## Introduction

According to Kellum et al., 2021, Acute kidney injury (AKI) affects approximately 30% of patients in the intensive care unit (ICU), and exhibits mortality rates of up to 50%. AKI is defined as a sudden loss of kidney function defined by a reduced urinary output or a rise in serum creatinine and is decomposed into four stages. Stage 1 (initiation) of AKI, where patients do not exhibit clinical signs. Stage 2 (oligo-anuria) describes the phase when urine output is diminished, which occurs in 70% of AKI events, which leads to the dysfunction of various organs due to uremia. Stage 3 (polyuria) is the delicate phase when recovery is ongoing, however loss of water together with electrolytes such as potassium and sodium can lead to clinical complications. Finally, stage 4 refers to the restitution of kidney function (Kellum et al., 2012). Early detection of kidney injury (in the preclinical stage 1 phase) is crucial for providing adequate treatment and preventing further kidney damage by carefully balancing fluids through either loop diuretics or fluid administration as well as withholding contrast material administration, hyperglycemia and other kidney stress-inducing drugs (Kellum et al., 2021). Current standards of care are insufficient for treatment because increases in serum creatinine lag substantially behind renal injury (Hermansen et al., 2021; Mårtensson, Martling, and Bell, 2012). Consequently, rule-based systems for AKI based on creatinine or other lagging molecular biomarkers have not been shown to improve clinical outcomes (Lachance et al., 2017).

Therefore, there is a need for accurate and personalized predictors able to anticipate AKI early in its development process. For similar prediction tasks, machine learning approaches have been shown to thrive in a data-rich environment such as the ICU. Examples include early circulatory (Hyland et al., 2020) and respiratory failure prediction (Hüser et al., 2024). A number of challenges need to be overcome for such a predictor to effectively support the workflow of ICU clinicians. Firstly, it needs to deliver timely, actionable and relevant alerts to doctors of possible AKI events to avoid alarm fatigue. Secondly, each patient should receive alerts based on their individual clinical profile. Thirdly, such an early warning system needs to generalize well to various patient demographics and healthcare systems, which has been proven to be particularly difficult in AKI occurring in the ICU specifically (Tomašev et al., 2019; Cao et al., 2022). Fourth, an early warning system deployed in a clinical context should motivate the predominant factors behind any given alarm, to aid decision making and increase adoption using, for instance, SHAP values (Lundberg and Lee, 2017) or other model inspection methods.

Several prediction models for AKI have been proposed in the literature, however they each come with important caveats. First, the performance of models solely based on serum creatinine *or* urine output do not make them clinically applicable due to the lack of timely sensitivity of those biomarkers (Alge and Arthur, 2015). A previous attempt at predicting AKI from electronic health records using ML failed to demonstrate adequate clinical performance (Mohamadlou et al., 2018). While a recent LSTM-based method (Tomašev et al., 2019) showed good performance, adequate population generalizability was lacking, and has since been explored to some extent by Cao et al., 2022. Another limitation of previous ML approaches, including Tomašev et al., 2019 and subsequent follow-up work by Cao et al., 2022, is that AKI events are only labeled using serum creatinine. This label definition does not fully conform to the definition of AKI as given by the KGIDO foundation, which requires both serum creatinine and urine output. Finally, the data used to train ML models, particularly the dataset of the United States Veterans Affairs used in the models of Tomašev et al., 2019 and revisited by Cao et al., 2022, are not publicly available. This conflicts with FAIR (Findable, Accessible, Interoperable, and Reusable) guidelines for scientific data sharing (Wilkinson et al., 2016) and limits the reproducibility of the previously reported work. In this work, we use open datasets for our analyses (HiRID-II and MIMIC-IV that will be or are available on PhysioNet.org) and make our code (https://github.com/ratschlab/AKI-EWS) open-source, to ensure the end-to-end reproducibility of our results and applications of our models on real world data.

Here, we present a detailed benchmark of gradient-boosted decision tree (GBDT) models, which have been specifically adapted to AKI prediction from multi-variate ICU time-series, and the LSTM-based model proposed by Tomasev. et al., which to our knowledge has not been specifically evaluated in ICU data sets. Our evaluation employs the complete KDIGO definition of AKI, incorporating both serum creatinine levels and urine output criteria. We evaluate model performance across a wide array of challenging machine learning situations arising when deploying models in a clinical setting. First, we examine the performance in two distinct health systems by assessing the performance of models in the HiRID-II (Switzerland) as well as the MIMIC-IV (United States) data sets. Second, we evaluate the fairness of both models in sub-cohorts stratified by age, gender, and diagnostic group category. Third, motivated by Tomašev et al., 2019, whose model was trained in a male-dominated US veteran cohort, we investigate gender transferrability by training each model on both genders separately, and then evaluating it on the other. Fourth, we investigate the performance of the two models in an artificially reduced data setting, which simulates low-resource health settings with a scarcity of available labeled data. Fifth, we investigate the clinical plausibility of the most important parameters identified by each of the methods, by comparing its agreement with relevance rating provides by experienced ICU clinicians. We conclude with a case-study on the impact of fluid and furosemide administration on model performance by estimating the treatment effect, inspecting SHAP values and quantifying the effect on false alarm rates.

In summary, our study makes significant advancements in the field of AKI prediction in ICU settings, with the following key contributions:

- Utilization of the complete KDIGO definition of AKI, incorporating both serum creatinine levels and urine output, to address existing gaps in predictive modeling.
- Development of a novel time-based stacking technique to improve GBDT AKI prediction performance.
- Training and evaluation of a previously proposed LSTM-based approach on an open ICU data set.
- Comprehensive assessment of model performance across different health systems and patient demographics, focusing on generalizability and fairness.
- Investigation of gender transferrability and model robustness in settings with limited data, highlighting the versatility of the proposed models.
- An explorative study of the effect of AKI treatments on model performance, with an investigation of its possible mechanism using propensity-matched treatment effect analysis and model introspection using SHAP values.
- Supporting reproducibility, demonstrated by the use of open datasets and the open-sourcing of our codebase.

## Methods

Figure 1 presents an overview of the data processing, modeling and evaluation pipeline. Below, we describe each element of the pipeline in detail.

**Figure 1.**
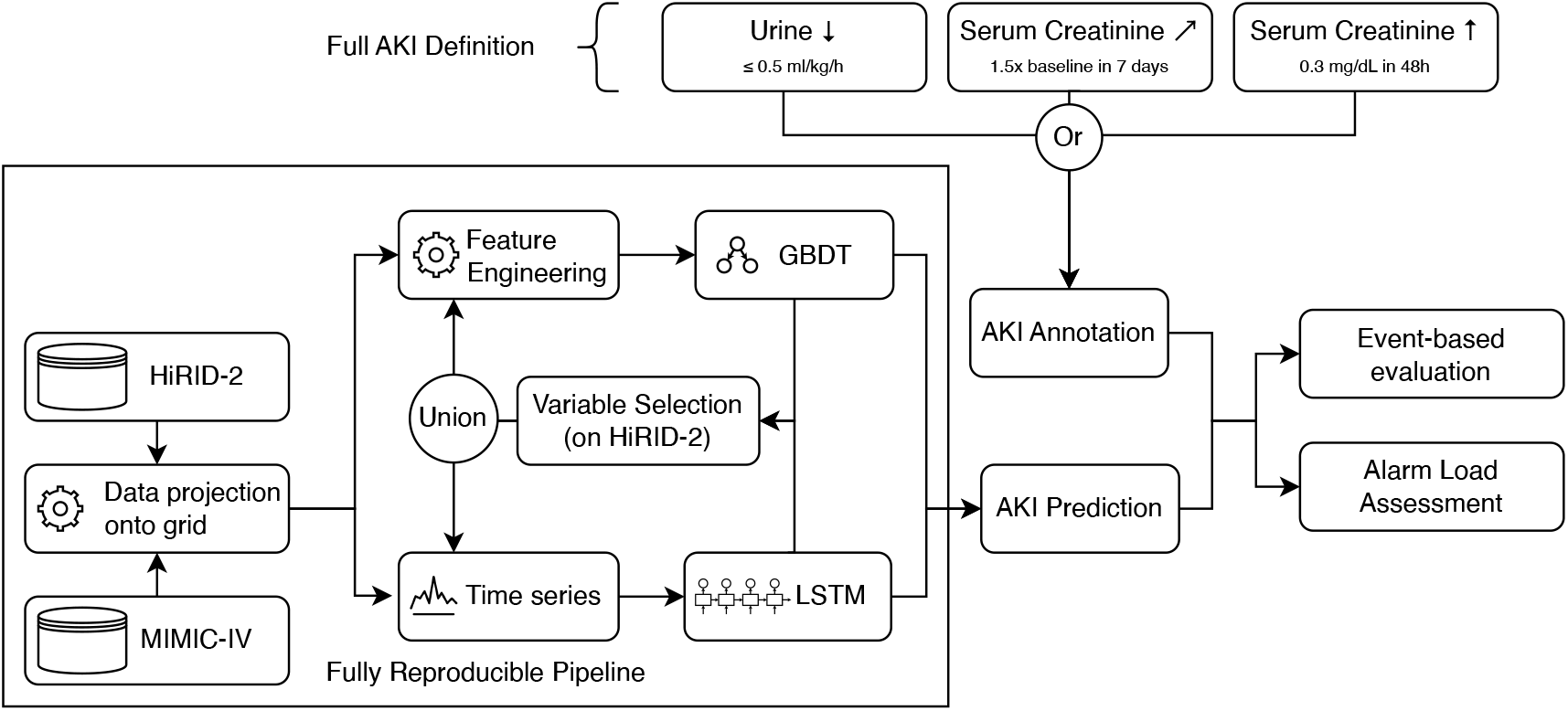
Overview of the machine learning and evaluation pipeline for AKI prediction in the ICU using the complete KDIGO definition. As our models are designed for ICU settings, and not the general ward, as often done in previous literature, where such measurements are not available, we can use the full KDIGO AKI definition, i.e. 1.5x increase in creatinine compared to a baseline measured in the last 7 days, a 0.3mg/dL creatinine increase within 48h or a reduced urine output below 0.5 ml/kg/h. With this definition, we built a reproducible machine learning pipeline aiming for the early identification of AKI. Two model types are used, one is the LSTM-based model proposed by Tomašev et al., 2019 which uses the time series directly, while the other uses gradient-boosted decision trees (GBDT) (Ke et al., 2017). Both models are built on a set of variables that are most important for inferring kidney failure determined using SHapeley Additive exPlanations (Lundberg and Lee, 2017) and greedy forward selection to maximize model portability in practice by reducing the number of required variables.

### Study Data Sets

The HiRID-II data set (Hüser et al., 2024) was used to test performance in the Swiss health system. The dataset was previously k-anonymized with respect to observable attributes including age, gender, weight and height and absolute date information was removed (Hüser et al., 2024). The MIMIC-IV data set (Goldberger et al., 2000; Johnson et al., 2023) was used to assess performance in the US health system. From both data sets patients were excluded from the study if their AKI status cannot be reliably annotated at any point of their ICU stay. The same processing pipeline including partial imputation, AKI annotation, and feature engineering was used for both data sets. Exploration of the prediction task and variable selection was guided by HiRID-II. The HiRID-II study data set contained 52,356 ICU admissions (363.92 patient years), while the MIMIC-IV study data set contained 45,221 ICU admissions (502.43 patient years).

### AKI Annotation using the KDIGO Definition

Acute Kidney Injury (AKI) is characterized by a decreased glomerular filtration rate (GFR), indicative of renal injury and impaired function (Kellum et al., 2012). In clinical practice, serum creatinine levels are often used as a proxy for GFR. This condition is classified into three stages, each indicating a progressively severe level of kidney dysfunction. In our study, we specifically examined the progression from a stable condition to any of the more severe stages (AKI 1-3), to include early, pre-clinical signs of deterioration. The criteria we used to identify the transition to stage 1 (AKI-1) or a higher stage are as follows:

- Increase in creatinine by ≥ 0.3 mg/dl (≥ 26.6 mmol/l) within 48*h*
- Increase in creatinine to ≥ 1.5 times baseline within 7 days
- Urine volume < 0.5 ml/kg/h for 6 h
- Initiation of renal replacement therapy

For each patient, we determined the baseline creatinine level based on the lowest value observed during the entire ICU stay, including the period prior to ICU admission. The AKI status was continuously monitored and classified as stable or in an AKI stage ≥ 1, on an hourly basis throughout the ICU stay, using the criteria mentioned above.

Using prior knowledge by ICU clinicians, we incorporated two post-processing steps in our analysis. Gaps of up to 24 hours between two AKI events were bridged, treating the patient as in an AKI state during these intervals. The purpose of adopting this approach was to maintain consistency in situations where sporadic data may indicate a temporary return of stability. Additionally, AKI events lasting less than 4 hours were excluded from our analysis, considering these brief fluctuations as likely non-significant or spurious transitions.

### Partial Imputation and Feature Engineering

As a first step, data was partially imputed on a time grid with step size 5 min. Variable-specific “active periods” were defined using prior clinical knowledge, and data was forward filled up to these active periods, in which the last measurement can be assumed to be a reliable estimate. Beyond this horizon or if the patient had no measurements for the variables, data was left as missing on the time grid. Features were then computed, which consisted of the following classes

- Current (partially-imputed) value of all variables, including time-since-admission.
- Multi-scale history, i.e. summary statistics over several horizons in the past.
- Measurement history, which capture how often previously and when a measurement was last observed.

For an exhaustive description of these features we refer to Hyland et al., 2020 and Hüser et al., 2024.

### Variable Selection

To compare model performance between different models in an unbiased way, each model selected important variables independently. For the GBDT models, the SHAP method was used to pre-filter the variables. For each of the 5 data splits, the top 30 variables according to SHAP mean absolute values in the validation set were found. The intersection of these 5 sets yielded 23 variables. Then greedy forward selection on this set guided by AUPRC on the validation set was performed in the 5 splits, and the first *k* variables were included such that the model’s was within 1 % of the optimal performance, as done in related works (Hyland et al., 2020; Hüser et al., 2024). The variable sets found in the 5 splits were then intersected, yielding 15 variables. The LSTM model selected variables by assessing AUPRC performance loss in the validation set when randomly permuting feature vectors of individual variables, described as Permutation Variable Importance Measure (PVIM) by Wei, Lu, and Song, 2015. The union of the important variables found for GBDT / LSTM was used for all further experiments, which yielded 28 variables overall.

### Model Setup

Gradient-boosted Decision Trees (GBDT)

Since GBDT models are not inherently made to deal with time series data, we investigated several alternatives to enable GBDTs to use multivariate time series available in the ICU.

- `GBDT-snapshot`: As a baseline a model was used which only views the current patient state at time *t*.
- `GBDT-history`: This model uses manually constructed features which summarize the historical state of the patient, by computing aggregation functions over the past time series, as well as other features which capture measurement intensity. Additionally the model views the current patient state at time *t*.
- `GBDT-time-stacked`: Motivated by our initial observation of the high relevance of the *time since admission* feature for the prediction task, we designed a model stacking schema, whereby each model is specialized to be used at a particular period of the ICU stay, e.g., one model is used only in the first hour of the ICU stay. In this way the prediction task is conditioned on properties of different phases of the ICU stay. As a final step, all models post-hoc calibrated using isotonic regression, using validation set, to encourage smooth prediction score trajectories. This model setup is illustrated in Figure 2.

**Figure 2.**
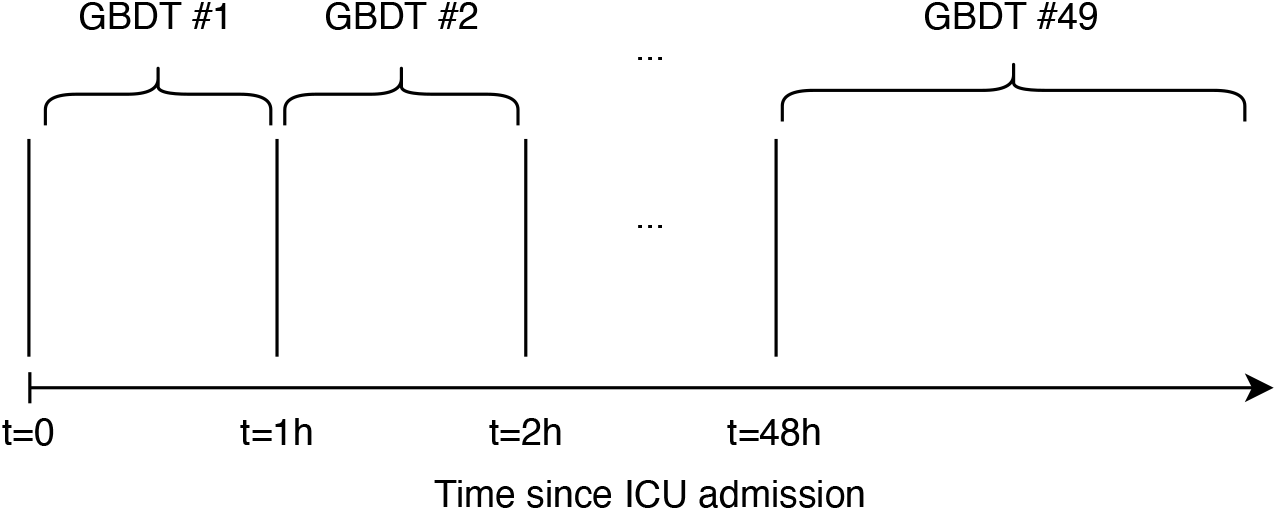
Stacked model setup. For each time period after admission, the AKI risk is estimated using a GBDT model trained on a window of time series that contain information only for that time period. The last model is used to estimate AKI risk on time-points beyond 48h after ICU admission. As postprocessing, all models are jointly calibrated using isotonic regression.

All GBDT models were implemented with LightGBM (Ke et al., 2017), using early stopping based on AUPRC performance in the validation set.

LSTM Model (Tomašev et al., 2019)

The same architecture as introduced by Tomašev et al., 2019 was kept. However, we reimplemented the model in Pytorch and changed some implementation details to accomodate the data sets and resolutions available in HiRID-II and MIMIC-IV. We provide more information on our implementation in Appendix 1.

### Data Splitting Setup

For HiRID-II, we used the provided split into development and test set (Hüser et al., 2024). The latter comprised the latest admissions of the HiRID-II dataset from June 2018-June 2019, to evaluate whether models generalize well into the future. The development data was split into a training set and a validation set at a 80%:20% ratio by complete ICU admissions, 5 times at random. The test set contained 3,753 patients.

For MIMIC-IV, since admission time is not provided, we cannot use the same strategy. First one fixed test set containing a random 10 % of ICU admissions was drawn. Then the remaining data was partitioned 5 times at random into a training set and validation set using a 80%:20% ratio. The test set contained 4,522 patients.

### AKI Event-based Evaluation

Given the predicted AKI risk every 5 minutes, we generate an alarm if the predicted risk score is above a threshold. We silence alarms for 8 hours after an alarm and do not produce alarms for 12 hours after an event. We used the same event-based evaluation scheme used by Hyland et al., 2020 that measures the fraction of identified AKI events, i.e., there was an alarm in the 24h before the event (event recall) and the fraction of true alarms, i.e., there was an event in the 24h after the alarm (alarm precision). This is done for multiple thresholds summarized in an alarm precision/event-recall curve.

### Assessment of Alarm Load for ICU Clinicians

To have a complete assessment of alarm load resulting from the early warning systems, we investigate each calendar day in all test set patient stays. We group the days by whether at least one AKI event will occur within the next 48 hours, i.e the next two days. All alarms raised in a patient calendar day without any AKI event in the next two days are false alarms. Alarms raised in a calendar day with at least one AKI event in the next two days are true alarms with very few exceptions. We assess models by the average number of alarms in both calendar day types. This measures how many alarms clinicians would expect from a patient who is likely to have AKI and how many false alarms they will expect from a patient who does not exhibit AKI. This evaluation is much more relevant to the clinical applicability of early warning systems than most previous works in the literature, who focus solely on ROC-based, and rarely on PR-based metrics.

## Results

### Creatinine-only AKI Definition is Restrictive in ICU Setting

In previous works on AKI prediction using ML methods, the full international consensus KDIGO definition of AKI was not applied. In particular, in the work of Tomasev at al., AKI was not specifically analyzed for ICU settings, and because of lack of continuous urine measurements, the full KDIGO definition could not be used. Our analysis in Table 1 shows substantial differences in the detection and characterization of AKI events between the official KDIGO definition vs. the proxy definition of Tomašev et al., 2019 and Cao et al., 2022. We observe that the full KDIGO definition results in a much higher prevalence of AKI events, with a 52% prevalence rate compared to 17% under the creatinine-only definition (3.06-fold increase). Furthermore, the number of detected AKI events using the KDIGO definition is 46,260, a 3.31-fold increase compared to the 13,974 events identified using the creatinine-only definition in the HiRID-II data set. This result casts doubts on whether a creatinine-only definition is sufficient for ICU settings, or does not miss most of the relevant AKI events.

**Table 1.** Comparison between the full KDIGO event definition and the serum creatinine-only definition used in Tomašev et al., 2019. The full definition shown in Figure 1 allows us to capture many more events, increasing the prevalence of AKI events in the HiRID-II dataset from 17% to 52%. The average duration of each event is shorter when using the full definition, indicating that considering urine leads to a more fine-grained AKI definition.

|  | Creatinine Only Definition | KDIGO Definition |
| --- | --- | --- |
| **Patients with at least one AKI event** | 10,522 (prevalence: 17%) | 31,999 (prevalence: 52%) |
| **Number of labeled AKI events** | 13,974 | 46,260 |
| **Total AKI hours** | 594,778 | 1,163,816 |
| **Average AKI event duration (in hours)** | 42.6 | 25.2 |

The KDIGO definition identified approximately 1.96 times more than the total AKI hours calculated under the creatinine-only definition (594,778 hours). However, the average duration of each AKI event was markedly shorter with the KDIGO definition (25.2 vs 42.6 hours) under the creatinine-only definition. This indicates that the KDIGO definition not only identifies more AKI events but also characterizes them with a finer granularity, resulting in a more refined and detailed understanding of the AKI event timeline.

Taken together, these findings underscore the importance of considering urine output in the definition of AKI, as it leads to a more comprehensive and nuanced detection of AKI events, enhancing the precision of clinical and epidemiological studies in this domain. Our following benchmark on AKI prediction in the ICU setting is to our knowledge the first one to take into account the full KDIGO definition.

### GBDT variants for AKI Prediction

Since the LSTM model introduced by Tomašev et al., 2019 is inherently time-dependent, we investigated how we could incorporate this time dependence in classical ML methods like GBDT. Two modifications were investigated here, and are described in detail in section. Table 2 compares the performance of “time-stacked” GBDT models for each hour after ICU admission compared to one model for all timepoints, as well as showing the performance of the GBDT model trained using multi-scale history features vs GBDT models trained using snapshot features, i.e. the current patient state only. We observe that the time-stacked models trained on data from each hour performs better than a single joint GBDT model trained on data from all timepoints. At 80% event recall, we observe that the time-stacked GBDT model with snapshot features performs slightly better than the time-stacked GBDT model trained on the multi-scale history features. In terms of AUPRC, the highest performing setup was the time-stacked GBDT model trained on each hour using the snap-shot features. Therefore, we proceeded to use this setup for the remainder of the analyses unless otherwise stated.

**Table 2.** Comparison of GBDT models modeling all time points in the ICU stay using one model vs one model for each of the 48h after ICU admission and one model for the remaining timepoints. Overall, 49 models were used, and then re-calibrated as explained in the stacking framework diagram. The LSTM-based model is used as a comparison. The performance results were obtained on the HiRID-II test set.

| Model type | LSTM | Joint GBDT |  | Time-stacked GBDT |  |
| --- | --- | --- | --- | --- | --- |
| Feature type | Time-series | Snapshot | History | Snapshot | History |
| Event-based AUPRC | 0.626 ± 0.003 | 0.641 ± 0.002 | 0.637 ± 0.001 | **0.657 ± 0.004** | 0.647 ± 0.005 |
| Alarm precision (%) @ Event recall=80% | **49.3 ± 0.9** | 47.3 ± 0.3 | 48.5 ± 0.4 | **49.2 ± 0.6** | **49.0 ± 0.5** |
| False Alarm Frequency (/patient/day) | 0.876 ± 0.279 | 0.973 ± 0.322 | 0.798 ± 0.017 | **0.742 ± 0.018** | 0.770 ± 0.015 |

### Performance of Time-stacked GBDT vs. LSTM in Different Health Systems (Switzerland & USA)

A good AKI predictor should have robust performance across different health systems. Here, we compare the performance of the best performing GBDT variant and LSTM on data collected from two health systems from different countries, namely HiRID-II from Switzerland and MIMIC-IV from the USA.

Table 2 and Figure 3 compare the performance of the LSTM model originally proposed by Tomašev et al., 2019 and our best performing GBDT-based variant on the HiRID-II data set. We observe in Figure 3a that the LSTM-based method achieves the same precision at 80% recall but on average lower AUPRC compared to the GBDT model. Figure 4b shows that the false alarm load of the GBDT model is slightly lower than that of the LSTM model.

**Figure 3.**
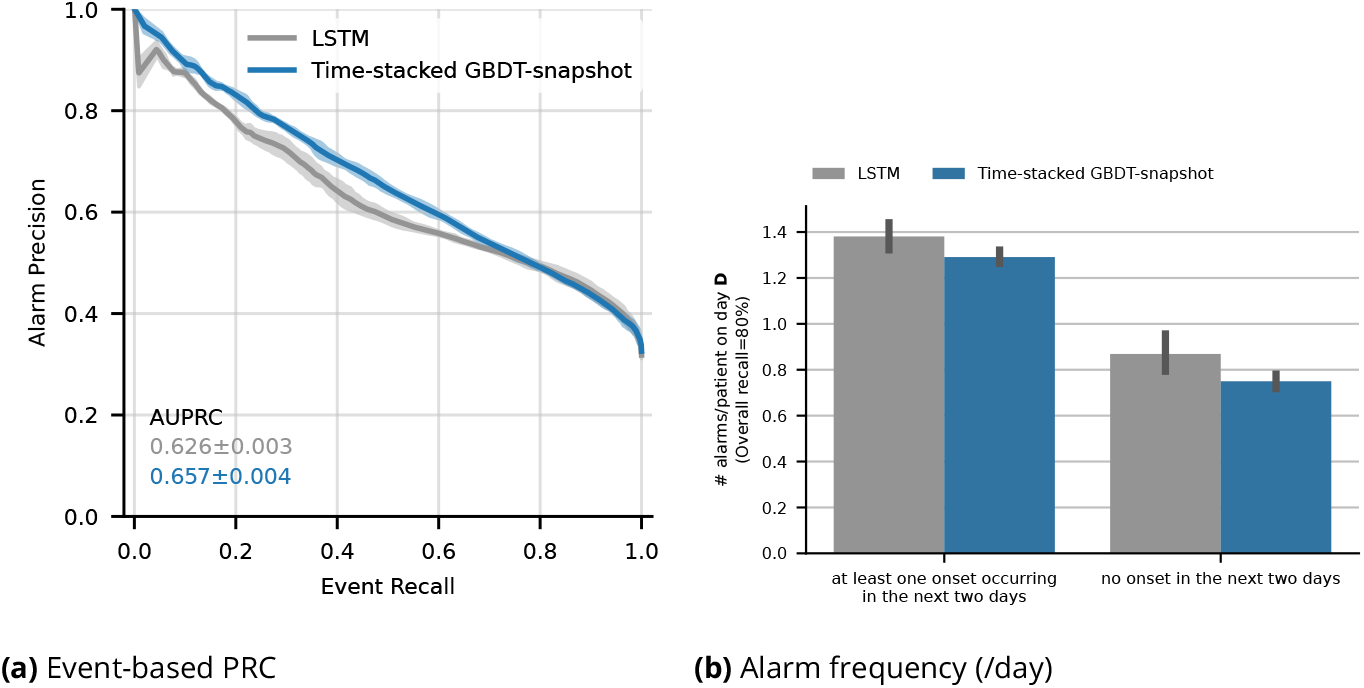
Comparative performance of our proposed time-stacked GBDT model versus the LSTM-based model proposed by (Tomašev et al., 2019) on the HiRID-II test set.

**Figure 4.**
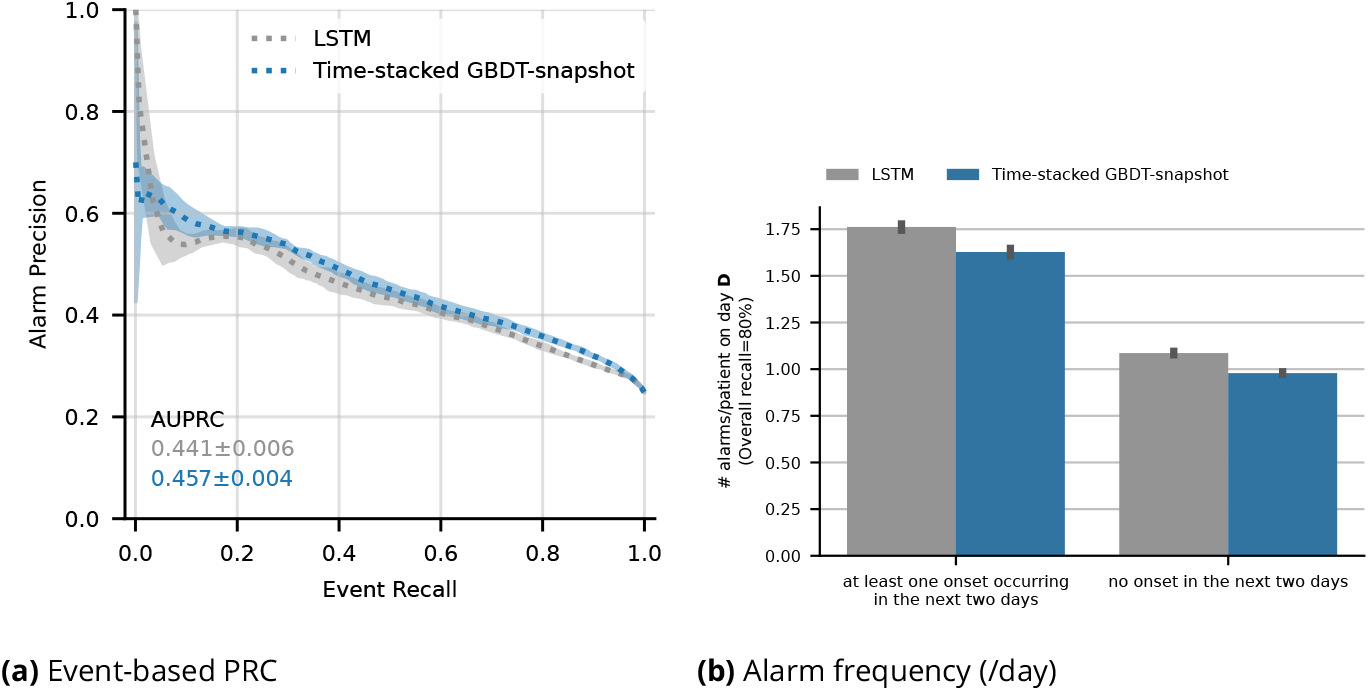
Comparative performance of time-stacked GBDT versus the LSTM-based model on the MIMIC-IV test set. Both models used 22 out of the 28 variables that were identified by the main variable selection procedure on HiRID-II. The remaining 6 variables could not be extracted from the MIMIC-IV data set.

Figure 4 compares the LSTM and the GBDT relative performance on the MIMIC-IV dataset, and we can see that both the AUPRC and the precision at 80% recall is higher for the GBDT model, similar to what we observed in the HiRID-II dataset in Section. The alarm load of the GBDT model is again slightly lower than that of the LSTM model when both models retrieve 80% the upcoming AKI events.

### Performance in the Limited Training Data Regime

Data scarcity often hampers the development and deployment of clinical machine learning models in practical settings (D’Hondt et al., 2022). As a result, we investigated the effect of reducing training set size on model performance for different modeling strategies, namely the GBDT-history and GBDT-snapshot models as well as the LSTM model^1^. The latter is a deep learning model, a class of models known to be data-intensive (Thorsen-Meyer et al., 2022). As we can see in Figure 5, the following trend holds down to about 5% of the training data: GBDT-snapshot outperforms GBDT-history, which outperforms the LSTM model. Under this threshold, error bars overlap significantly, and all models exhibit high variance across temporal splits and are hard to distinguish in terms of performance on the same test set. This study confirms that under data constraints, the GBDT-snapshot model is more robust compared to GBDT-history and LSTM until the training dataset is reduced to less than 5%, beyond which point all models show similar levels of high variability. It is also noteworthy that AUPRC performance loss is relatively minor, even at extremely small training set sizes, corresponding to only 40 patients at 0.1 % training set size.

**Figure 5.**
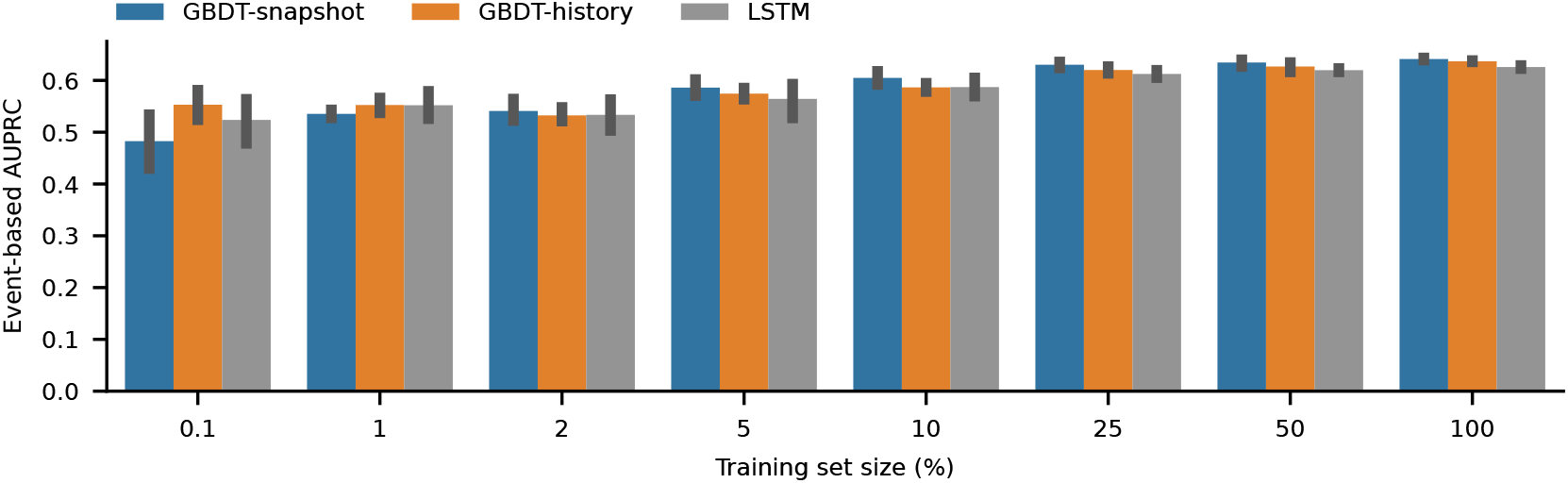
Performance of GBDT-snapshot, GBDT-history, and LSTM models when progressively increasing fractions of the original training set are used to train each model. The smallest training set contained only approximately 40 patients. This analysis was performed on the HiRID-II test set.

### Fairness of AKI Predictors Analyzed through Sub-cohort Analyses and Domain Transfer

As highlighted by Cao et al., 2022, the previous state-of-the-art model proposed by Tomašev et al., 2019 was initially analyzed on a male-dominated (94%) patient cohort of the United States Veteran’s Affairs clinical dataset, substantially hampering the ability to determine the performance of the model on females. While Cao et al., 2022 have addressed this, we also wanted to assess both GBDT-and LSTM-based models performances on sub-cohorts divided by gender, age, and diagnostic category. We evaluate this in two ways: first, we analyze the performance of each model on each sub-cohort in the test set. Additionally, we evaluate the ability of the model to transfer from one gender to the other (e.g., from male to female and vice-versa). We observe that the GBDT-based model performs better in all investigated sub-cohorts (Figure 6). Both models show fair performance mostly, with the exception of patients in the age group 16-30, which exhibit reduced performance. As a side observation, due to the AKI prevalence differences across different sub-cohorts, the original event-based AUPRC values cannot fully reflect whether a model performs fairly for each sub-cohort. Therefore we corrected the prevalence of all sub-cohorts to be similar to that of the entire population using the false alarm scaling factor proposed by Hyland et al., 2020. After correcting for the different AKI prevalences, neither of the models exhibit issues with model transfer between genders. The LSTM-based model exhibits slightly larger variance in the different splits (Figure 7).

**Figure 6.**
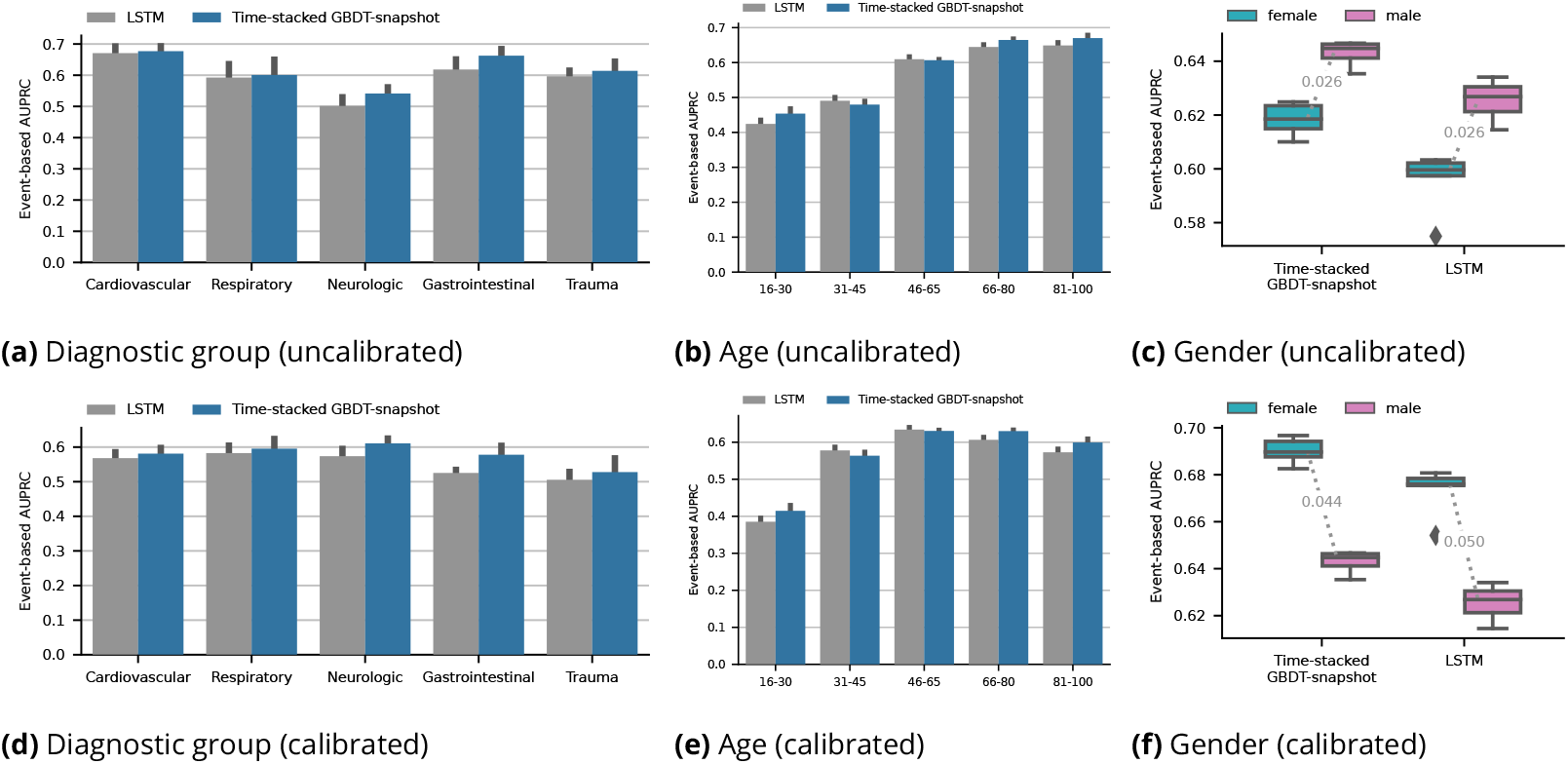
Performance in sub-cohorts of the HiRID-II test set. The first row of plots shows uncalibrated results, the second rows shows prevalence-corrected results, to eliminate the effect of different AKI prevalences in the sub-groups on the performance comparison.

**Figure 7.**
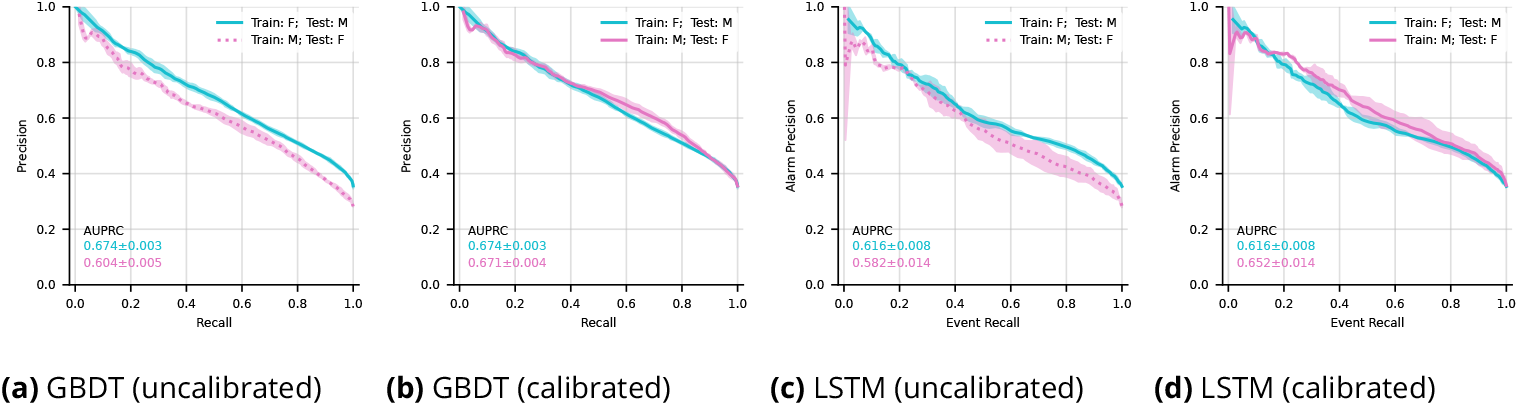
Gender transfer performance for a GBDT-based model and LSTM on the HiRID-II test set. (**F** and **M** refer to Female and Male respectively.). The left panels show uncalibrated performance results, and the right panels show results where the precision was corrected for AKI prevalence differences in the two genders.

### Clinical Plausibility and Feature Importance

We aimed to identify the key clinical parameters driving model performance as well as investigate the agreement of those features with clinician assessments. Figure 8a shows the importance of each variable by measuring the difference in performance in terms of AUPRC of the model trained normally and a model with individual variables corrupted. The GBDT-based model shows the strongest dependency on ‘time since admission’; enteral feeding and weight come as equally important features in second place. This strong reliance on ‘time since admission’ could partially explain the success of the time-stacking strategy. For the LSTM model, the model shows the strongest reliance on dialysis, which intuitively make sense, as dialysis is commonly associated with various stages of AKI (Kellum et al., 2021). Enteral feeding and urine output as well as C-reactive protein, EtCO_2_ and creatinine were also considered important. The concordance of important variables with those annotated as relevant by clinicians is shown in Figure 8b. For small model sizes, the variables selected by the LSTM are more clinically plausible, however this effect diminishes for medium-sized models, i.e., d=15. Then the GBDT-based model shows moderate clinical plausibility with a mean agreement score of 2, surpassing the LSTM-based model.

**Figure 8.**
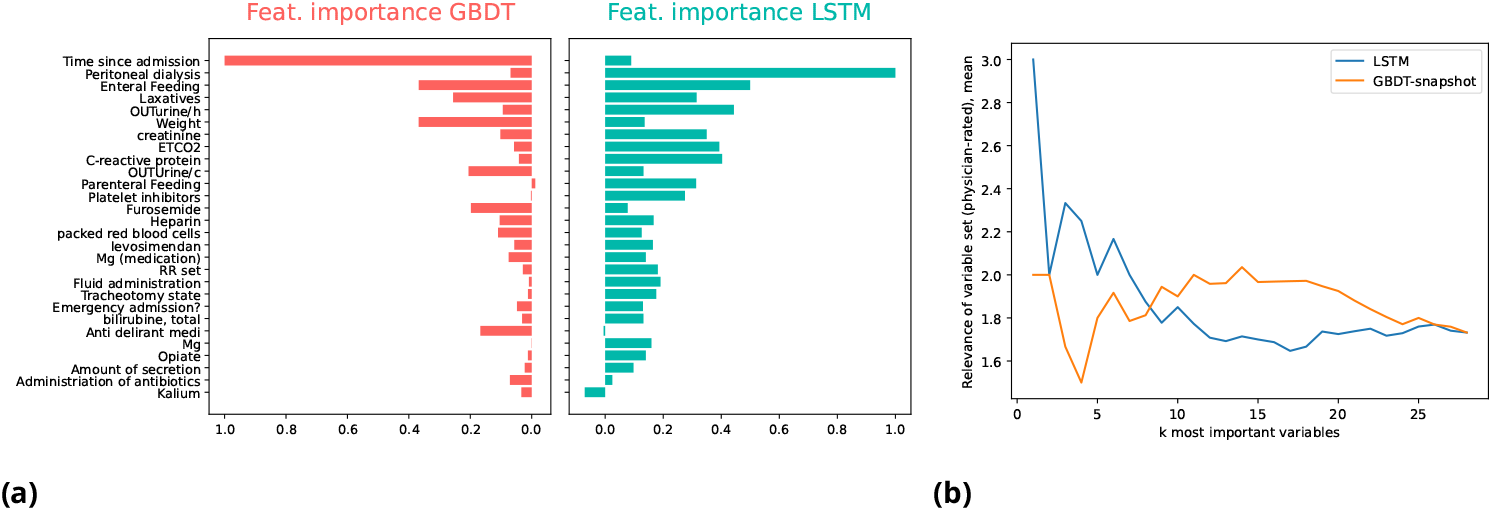
Variable importance inspection of the LSTM and GBDT-based model and comparison with ICU clinician provided relevance ratings. The HiRID-II validation set was used to retrieve the variable rankings. **(a)**. Feature importance of individual variables for each model. The importance was measured by shuffling the feature column at random, and measuring the resulting AUPRC performance loss on the validation set. The variables are ordered on the y-axis by the mean importance across the 2 models. **(b)**. Agreement between model-derived variable relevance and clinician relevance of the *k* most important variables for each model, as measured by mean annotated relevance manually assigned by two ICU physicians.

### Effect of Fluid/Furosemide Treatment on Model Performance

A property of our model is that it is trained on a data set of patients that received intensive medical care - hence there are no untreated controls, and patients may even have received treatments that affect the onset or occurrence of the event we would like to predict. AKI is a treatable condition, and fluid or furosemide (a loop diuretic) treatment can be used to manage the condition (Kellum et al., 2021). To explore their effect on our predictions, we investigated the interplay of treatments and predictive performance of the GBDT model. We observe that the prevalence of AKI among patients with any of the two treatments is elevated (Table 3). Patients with any of the two treatments show a 2.1% increase in false alarm rate, suggesting a slight loss in prediction precision. In Figure 9a) a treatment-effect analysis of fluid/furosemide on occurrence of AKI in the next 60h is shown. Time points where fluid/furosemide were given were propensity-matched with control time-points with equal AKI risk according to a GBDT-based predictor (but without using Fluid and Furosemide as input, respectively). We observe that both treatments slightly reduce future AKI risk. SHAP value inspection (Figure 9b/c), however, showed that the GBDT model associated fluid/furosemide administration with higher predicted risk of AKI. We hypothesize that the model could not correctly model the causal effect on AKI outcome, resulting in higher false alarm rates.

**Table 3.** Effect of medical treatments, namely furosemide and fluid input, on the false alarm rate of the GBDT-snapshot model. Performance is shown on the HiRID-II test set which was divided into two sub-cohorts.

| Sub-cohort | AKI Event Prevalence (%) | False Alarm Rate (%) |
| --- | --- | --- |
| Patients w/o fluid or furosemide treatment | 22.56 | 51.2 ± 0.95 |
| Patients with fluid or furosemide treatment | 33.06 | 53.3 ± 0.65 (2.1%↑) |
| *p*-value | - | 0.008 |

**Figure 9.**
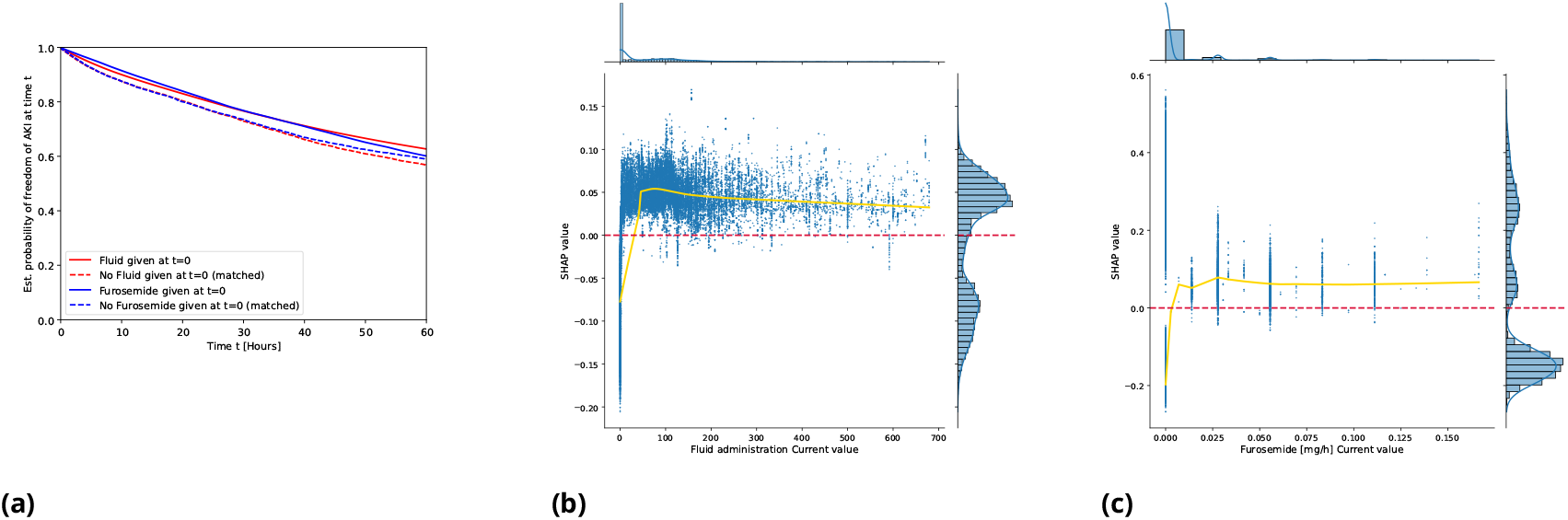
**(a)**. Treatment effect analysis of the effect of fluid/furosemide administration on medium term AKI risk (conditional on current freedom from AKI). Freedom of AKI curves of time points at which furosemide or fluids were given vs. propensity-matched controls time-points. t=0 denotes the time at which the treatment was initially given. A Kaplan-Meier estimator was used to estimate survival curves from right-censored data. **(b)/(c)**. SHAP analysis of relevance of fluid/furosemide values for predictions of the GDBT-snapshot model. Values above the red dotted horizontal line denote SHAP values which indicate a perceived AKI risk increase by the model, below a perceived decrease, respectively.

## Discussion

Our study on AKI prediction in ICU settings across two different health systems has addressed critical shortcomings in predictive modeling of AKI by using the complete KDIGO definition, incorporating both creatinine levels and urine output, in contrast to previous studies. We show that an incomplete definition would severely underestimate the prevalence of AKI events in an ICU context, and misses a significant portion of AKI events.

Additionally, we have developed a novel time-stacked gradient-boosted decision tree (GBDT) model that uses only 28 features and a time-since-admission stacking approach. This strategy represents a departure from the extensive manual feature engineering that was commonly employed in prior research for AKI prediction. For example Tomašev et al., 2019 use a total of 3,599 features for their baseline gradient-boosted trees model. This simplifies the process of feature extraction and reduces implementation complexity. Our approach was motivated by the high relevance of ‘Time-since-admission’ for AKI prediction in GBDT-based models. Future research should investigate the reasons behind the relevance of this feature and why the stacking approach actually helps.

In our comprehensive evaluation, the LSTM model proposed by Tomašev et al., 2019 was assessed in parallel with the GBDT model across two different health systems and patient demographics. This extensive analysis has confirmed the generalizability and fairness of both predictive models. Moreover, our evaluation of gender transferability and the robustness of the models in limited data settings has revealed their adaptability to a wide spectrum of clinical contexts, with robust performance down to a training set size of only around 40 patients. Observed performance differences between LSTM-based and the GBDT-based models were minor across settings, with GBDT, however, consistently outperforming the LSTM-based model. Both models show a high degree of fairness across sub-cohorts, and gender transfer did not pose an issue to both models in our experiments. This stands in contrast with the previous work by Tomašev et al., 2019, who observed issues in using models trained on male-only cohorts in female cohorts, however, for non ICU-specific settings (Cao et al., 2022).

Our study corroborates previous research findings which suggest that gradient-boosted decision trees (GBDT) exhibit superior performance on EHR-derived tabular data compared to deep learning (DL) methods (Yeche et al., 2021; Grinsztajn, Oyallon, and Varoquaux, 2022). The superior performance of GBDT in our study, especially in the context of AKI prediction, underlines the potential efficacy of this method on complex, multidimensional medical data. This is further amplified when adopting modeling approaches tailored to the prediction problem, such as time-based stacking.

Lastly, our commitment to reproducibility is evidenced by use of open datasets such as HiRID-II and MIMIC-IV. This, we hope, promotes further exploration and application of our work within the broader scientific community.

## Conclusion

This empirical study represents a substantial step forward towards precise early prediction of AKI in ICU settings, by employing the full KDIGO-based definition of AKI events. One of our most important observations was the impact of neglecting this complete definition, which ignored most of the relevant AKI events in our data set. Furthermore we conducted an extensive benchmark of two machine learning methods using (1) gradient-boosted decision trees as well as (2) an existing LSTM model from the literature. We observed that GBDT-based methods based on a time-based stacking schema outperformed the LSTM model. Both models performed robustly across a wide range of settings relevant in the ICU, including sub-cohort fairness, performance in the limited training data regime, and gender transfer. Future work will aim to reproduce and further the findings of our work, e.g., through a prospective validation of the early warning system or through coupling our predictions with early warning systems for other medical complications.

## Data Availability

Data availability statement:
Data used in this study were obtained from University Hospital Bern and we cite Hüser et al. for its use (https://www.medrxiv.org/content/10.1101/2024.01.23.24301516v1).

## Appendix 1

### Re-implementation of the LSTM-based model by Tomasev et al

Here we present an in-depth discussion of our LSTM model, a modernized reimplementation of the original model described by Tomašev et al., 2019. Our version, developed using PyTorch, includes several key modifications to enhance its performance and capabilities.

The decision to reimplement the LSTM model in PyTorch was driven by several factors. Primarily, the original implementation was based on TensorFlow 1.X, which was superseded in 2019 and is less readable compared to more modern frameworks like Tensorflow 2.X or Pytorch 2.X. This change to PyTorch 2 not only enhances readability but also ensures the longevity and maintainability of the code. Additionally, the GitHub repository for the original model has been archived, suggesting a lack of ongoing support and updates for the codebase. Another significant factor in our decision was the data representation format. The original model used protocol buffers to represent patients’ medical records, a format that is not widely adopted in the machine learning community. In contrast, our implementation uses torch tensors or numpy arrays, aligning with standard practices in the field.

Our model exhibits several key differences from the original implementation by Tomašev et al., 2019. In terms of model structure, while the original implementation includes a cumulative distribution function layer accommodating multiple prediction horizons (see Extended Data Fig. 2 of Tomašev et al., 2019), our model is tailored to predictions for a single horizon. Preliminary experiments indicated that the inclusion of multiple horizons did not improve the prediction performance.

Regarding feature engineering, there are noteworthy differences. The original model aggregated data into a 6-hour grid, incorporating a variety of summary statistics within each interval. In contrast, our model uses simpler features without such aggregation, owing to the 1-hour resolution data available in the MIMIC-IV data set. Furthermore, Tomašev et al., 2019 included the construction of high-level features, like the occurrence of groups of variables within each 6-hour bucket. Our implementation does not follow this approach, primarily because our data set consists of only 28 variables, which simplifies the process of feature engineering. In addition to the 28 variables, we also constructed binary presence indicators for all variables at each time step, to enable our models to distinguish between the absence of a numerical value and an actual value of zero. Regarding the auxiliary prediction task of predicting the maximum future observed value of a set of variables over the same horizon that we used to make the future AKI predictions (i.e., 48 hours), we selected the following target variables guided by ICU clinicians: serum creatinine, accumulated urine output, fluid input and loop diuretics.

## Footnotes

1 Time-stacked GBDT, our best-performing model as seen in Table 2, is not feasible in this setting, due to the lack of training samples to train 49 models, for very small training sets.

